# Failing maternal-fetal tolerance in Systemic Lupus Erythematosus (FaMaLE): a prospective cohort study for finding the molecular mechanisms behind pregnancy complications

**DOI:** 10.1101/2025.01.30.25321383

**Authors:** W. Dankers, J.F. van Ruitenbeek, S.A. Germe, A.R. Parra Sanchez, M.F.H.M. van Gaal, K. Cramer, D.C. Rohrich-Heldens, M.A. de Boer, L.G.M. van Baarsen, I.E.M. Bultink

## Abstract

**Introduction:** Pregnant women with systemic lupus erythematosus (SLE) have an increased risk of maternal complications and adverse fetal outcomes. These include preeclampsia, preterm birth and fetal growth restriction. Interestingly, this increased risk persists in subsequent pregnancies, whereas it decreases in healthy women due to the development of maternal-fetal tolerance. As maternal-fetal tolerance is crucial for a healthy pregnancy, we hypothesize that its failure contributes to the increased risk of pregnancy complications in women with SLE. Therefore, we initiated the FaMaLE study to investigate the failure of maternal-fetal tolerance in pregnant women with SLE.

**Methods and analysis:** In the FaMaLE study, women with SLE and healthy women are included in their first trimester of pregnancy (< 14 weeks gestational age (GA)) at Amsterdam UMC. Throughout the pregnancy, data on SLE disease activity, pregnancy course, and medication use are collected. Peripheral blood is collected once per trimester, within 48 hours before delivery and 5-12 weeks post-partum. In addition, the placenta is collected after delivery. Whole blood, peripheral blood mononuclear cells (PBMC) and placenta samples are freshly analyzed by flow cytometry to assess immune cell composition. The resulting data are analyzed in relation to SLE disease course, pregnancy course and pregnancy outcomes.

**Ethics and dissemination:** The study has been approved by the Amsterdam UMC Medical Ethics Committee and all participating women will be asked to provide informed consent. The findings will be disseminated through peer-reviewed publications, presentations at scientific meetings and via patient organizations.

**STRENGTHS AND WEAKNESSES:**

- A unique prospective longitudinal study design, featuring the collection of serum, plasma and PBMC throughout and after pregnancy, alongside placental cells and biopsies from the same participants. This is complemented by detailed clinical data on SLE disease course and pregnancy course, and outcomes.
- Fresh flow cytometry analyses allow immediate assessment of cell composition in blood and placenta, without freeze/thawing effects
- The study does not include pre-pregnancy collection of serum, plasma and PBMC; however detailed clinical data are collected during this period.

## INTRODUCTION

Systemic lupus erythematosus (SLE) is a chronic autoimmune disease with the potential to affect all organ systems in the body. It is characterized by a disruption in immunological tolerance, leading to the production of autoantibodies targeting nuclear self-antigens^1,2^. SLE affects 1 in 1000 people worldwide, with a higher prevalence among individuals of non-Caucasian descent, and predominantly affects women during their childbearing years^1^. Unfortunately, up to 50% of pregnant women with SLE experience complications, such as pre-eclampsia, fetal growth restriction and preterm birth^3,4^. Importantly, almost 50% of preterm births have a spontaneous onset, the other preterm births are induced mostly because of pre-eclampsia or fetal growth restriction^5^. Partially due to (the increased risk of) pregnancy complications, 60% of SLE patients have fewer children than desired^6,7^.

Despite the significant impact on the health and wellbeing of pregnant women with SLE and their children, the exact underlying cellular and molecular mechanisms driving these complications are still poorly understood. Understanding of these biological processes is crucial to enable prediction, early detection and novel treatments for placenta-related pregnancy complications in SLE.

One of the requirements for a healthy pregnancy is a well-functioning maternal immune system, which facilitates proper placentation, triggers the labor process at the end of pregnancy, and ensures that the semi-allogeneic fetus is not immunologically rejected during pregnancy^8^. This latter phenomenon is called maternal-fetal tolerance, and is thought to be the reason why generally the risk of pregnancy complications decreases in subsequent pregnancies with the same father. Interestingly, this protective effect is absent in women with SLE^3^, suggesting that impaired maternal-fetal tolerance may play a central role in the increased risk for pregnancy complications in women with SLE.

Previous data suggest an altered immune response in pregnant women with SLE who experience pregnancy complications^9^. Furthermore, there are some studies reporting an increased deposition of neutrophils extracellular traps (NETs) and higher numbers of natural killer (NK) cells and extravillous trophoblasts in the placentas of women with SLE. However, if and how this dysregulation contributes to pregnancy complications in women with SLE is currently unknown. Therefore, to address these knowledge gaps, we have initiated the FaMaLE study in which we aim to delineate the differences in cellular composition and function at the maternal-fetal interface in women with SLE, both with and without pregnancy complications, compared to healthy pregnant women.

## METHODS AND ANALYSIS

### Study overview and design

This single-center prospective cohort study was initiated in May 2022 and will include 60 pregnant SLE patients and 18 healthy pregnant controls, all aged 18 years or older, who receive their antenatal care from the first trimester onwards at Amsterdam UMC. Women with SLE must be diagnosed according to the ACR 1997, SLICC and/or ACR/EULAR 2019 criteria and must be under treatment for their disease at Amsterdam UMC. The treating gynaecologist and rheumatologist will screen potentially suitable participants during their first trimester of pregnancy at the Department of Obstetrics and Gynaecology and the Department of Rheumatology and Clinical Immunology of Amsterdam UMC. Eligible subjects will be informed about the aim, procedures and potential risk of the study by members of the research team. Participants can be enrolled before 14 weeks gestational age after providing informed consent. An overview of the study protocol is shown in ***figure 1***.

**Figure 1.**
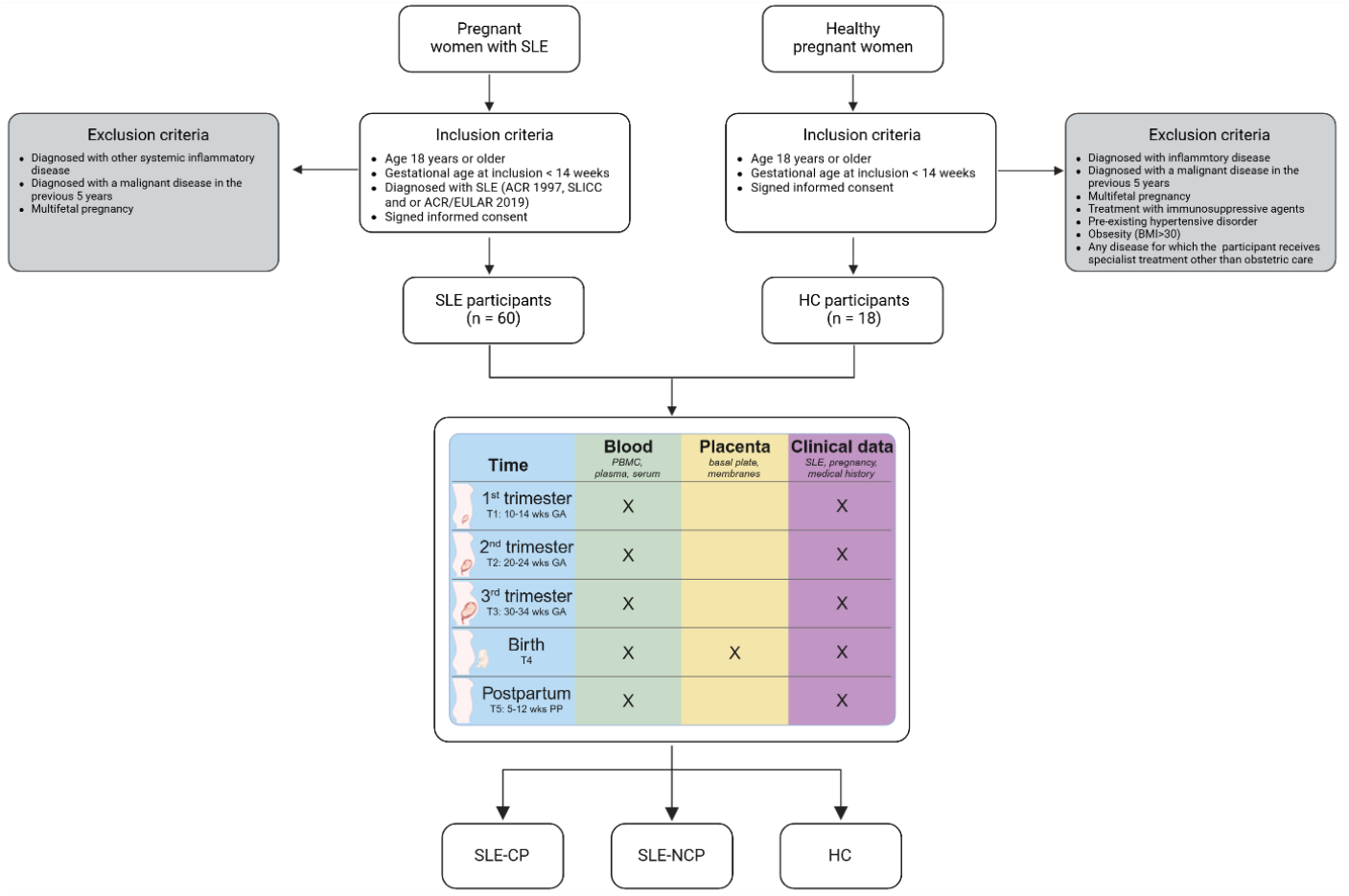
FaMaLE study flowchart. Abbreviations: SLE-CP, SLE complicated pregnancy; SLE-NCP, SLE non-complicated pregnancy; HC, healthy control. Created in BioRender. Dankers, W. (2025). https://BioRender.com/t19u876

Data collection includes demographic information (age, ethnicity, highest level of education), medical history (smoking status, alcohol/drug use, general and SLE-specific medical history, current and prior medication use) and obstetric history (number of previous pregnancies, any previous miscarriages, course and outcome of previous pregnancies). SLE disease activity will be assessed using the SLEPDAI during pregnancy^10^, and SLEDAI-2K^11^ before pregnancy and after delivery. Preconceptional disease-related damage is assessed by the Systemic Lupus International Collaborating Clinics/American College of Rheumatology (SLICC/ACR) Damage Index^12^. Additionally, gestational age at delivery and gender, weight and delivery outcomes for the newborn will be recorded. Apart from routine laboratory investigations for regular clinical care in pregnant women with SLE, at each visit an additional 45 ml of blood is collected in 4 heparin tubes and 1 coagulation tube. From these serum, plasma and PBMC are isolated. The blood sample from time point 4 (T4) is collected within 48 hours before delivery, when the woman is admitted at the delivery room. After delivery, the placenta will be collected for further analysis.

Medical data will be recorded by the treating gynaecologist and rheumatologist. Histopathological examination of all placentas will be performed by one assigned pathologist according to the Amsterdam Placental Workshop Group Consensus Statement^13^. Pregnancies are classified as complicated if they meet any of the following criteria: preterm delivery (<37 weeks gestational age), pregnancy induced hypertension (*de novo* or superimposed), pre-eclampsia (ISSHP criteria 2018^14^), small for gestational age child (<p10), placental abruption and/or intrauterine fetal death (>16 weeks gestational age).

### Inclusion and exclusion criteria

#### Inclusion criteria for SLE patients

- Age 18 years or older
- Gestational age at inclusion <14 weeks
- Antenatal care from first trimester onwards in the Department of Obstetrics and Gynaecology of Amsterdam UMC
- Diagnosed with SLE, fulfilling ACR 1997, SLICC and/or ACR/EULAR 2019 criteria
- Signed informed consent

#### Inclusion criteria for healthy pregnant women

- Age 18 years or older
- Gestational age at inclusion <14 weeks
- Antenatal care and delivery in the Department of Obstetrics and Gynaecology of Amsterdam UMC
- Signed informed consent

#### Exclusion criteria for SLE patients

- Diagnosed with a systemic inflammatory disease other than SLE
- Diagnosed with a malignant disease in the previous 5 years, except for basal cell or squamous epithelial cell skin cancers
- Multifetal pregnancy

#### Exclusion criteria for healthy pregnant women

- Diagnosed with a systemic inflammatory disease
- Diagnosed with a malignant disease in the previous 5 years, except for basal cell or squamous epithelial cell skin cancers
- Multifetal pregnancy
- Treatment with immunosuppressive agents
- Pre-existing hypertensive disorder
- Obstetric history of hypertensive disorder of pregnancy, fetal growth restriction (birth weight < p10), preterm birth, placental abruption or intra-uterine fetal death
- Obesity (BMI > 30)
- Any disease for which the woman receives specialist treatment other than obstetric care

### Sample size calculation

The rate of pregnant SLE patients to be included in this study is expected to be 15-20 per year based on our experience over the last >15 years in the multidisciplinary SLE pregnancy care team in Amsterdam UMC. Pregnancy complications due to placental insufficiency as defined above are expected in 30% of SLE patients as previously published by our research group^3^. In the present study women are categorized into 3 groups; healthy control (HC), SLE complicated pregnancy (SLE-CP), or SLE not complicated pregnancy (SLE-NCP).

In the analysis, differences in cell composition in whole blood, PBMC and placental tissue between these groups will be compared. We aim to detect differences greater than 10%, so at least 16 patients per group are needed for 80% power to detect differences (with an α of 0.05). Accounting for a 10% dropout rate throughout this study, 18 subjects per group need to be included. Considering an expected 30% incidence of pregnancy complications in SLE patients, 60 pregnant women with SLE are needed to include at least 18 patients with complications. Additionally 18 healthy pregnant women will be included.

### Study aims

The primary aims of this study are (1) to compare the cellular profile of placental cells from women in the HC, SLE-CP, SLE-NCP groups; (e.g. surface markers, transcription factors, gene expression), (2) to analyze the cellular function, cell-cell interactions and the development of tolerance at the *in vitro* maternal-fetal interface, and (3) to identify potential peripheral blood biomarkers associated with pregnancy complications.

As secondary objectives, the study aims to establish an *in vitro* organoid-based model for the maternal-fetal interface in SLE and to compare the cellular profile and different proteins in the peripheral blood of women in the three groups.

### Study procedures

#### Blood sampling

From the blood drawn at the five timepoints throughout pregnancy, serum and plasma will be isolated within four hours after blood draw from anti-coagulation and heparin tubes, respectively. They are stored at −80°C for later use for detection of biologic markers (antibodies, cytokines, (disease-related) proteins, cell-free RNA). 100 µl of whole blood is saved for analysis using flow cytometry, and then peripheral blood mononuclear cells (PBMC) are isolated within 24 hours from the heparin tubes, using Lymphoprep^TM^ (Serumwerk, Bernburg, Germany) density gradient centrifugation. 2-4 million PBMC are analyzed using flow cytometry, and remaining PBMC are frozen per 5-10 million cells in freezing medium (10% DMSO (VWR, Amsterdam, The Netherlands) and 90% Iscove’s Modified Dulbecco’s Medium (IMDM; Gibco) supplemented with 30% FCS, 0.05mg/ml gentamicin (Gibco) and 0.00036 (v/v)% β-mercaptoethanol.

#### Placenta sampling

After delivery the placenta will be collected by the research team and stored in 0,9% NaCl at 4°C until processing, which starts within 2 hours after delivery. Since most current protocols for processing placenta focus on obtaining either immune cells or trophoblasts, a customized protocol was used to capture both cell types based on previously published protocols^15,16^. The umbilical cord is removed from the placenta, leaving the insertion intact. 4 cm of umbilical cord is stored in formalin for histopathological examination and classification by the assigned pathologist, as well as 1/3 of the chorioamniotic membranes. The remaining decidua parietalis and chorion (PC) are separated from the amnion, cut off and placed in 1x PBS. Once the membranes and umbilical cord are removed, the placenta weight is determined. 3 biopsies from random locations at the placenta are taken using an 8-mm biopsy punch, covered in O.C.T., snap-frozen in liquid nitrogen and stored at −80°C. The placenta is then cut into slices of ~1-1.5 cm thick, to assess the percentage of calcifications throughout the tissue. At least two blocks of 2×1 cm, of which one includes the umbilical cord insertion site, and both blocks covering the entire depth of the placenta, are harvested and stored in formalin for histopathological examination and classification by the assigned pathologist. Subsequently, the basal plate (BP; decidua basalis and ~5mm of villi) is dissected and put in PBS. Now, both PC and BP are cut into small fragments (~2-3 mm diameter) and washed with 1x PBS until the PBS is clear. After washing with Roswell Park Memorial Institute (RPMI) medium, tissues are transferred to C-tubes (Miltenyi Biotec, Bergisch Gladbach, Germany) and resuspended in digestion mix (200mg/ml collagenase and 10mg/ml DNase (Roche) in RPMI). Tissue is then dissociated on GentleMACS (BP: 1x m-heart-02.01, PC: 2x h-tumor-02.01) and left to digest for 1 hour at 37°C in a shaking water bath. After 30 minutes there is another round of dissociation (all: 1x m-heart-02.01). Digested tissues are filtered through 100µm cell strainers (Miltenyi Biotec), after which red blood cells (RBC) are lysed using 1x RBC lysis buffer (BioLegend). Finally, cells are filtered through a 70µm cell strainer, after which ~5 million cells are analyzed using flow cytometry, 1 million cells are plated for subsequent culture and remaining cells are frozen as described for PBMC.

#### Flow cytometry

2-4 million PBMC, 5 million cells from BP and PC, and 100 µl whole blood are freshly analyzed for cellular composition using five flow cytometry panels: myeloid cells, B cells, T cells, ILC and non-immune cells (***Table 1-5***). With each sample, a reference control sample is taken along to account for batch effects during analysis.

**Table 1.**
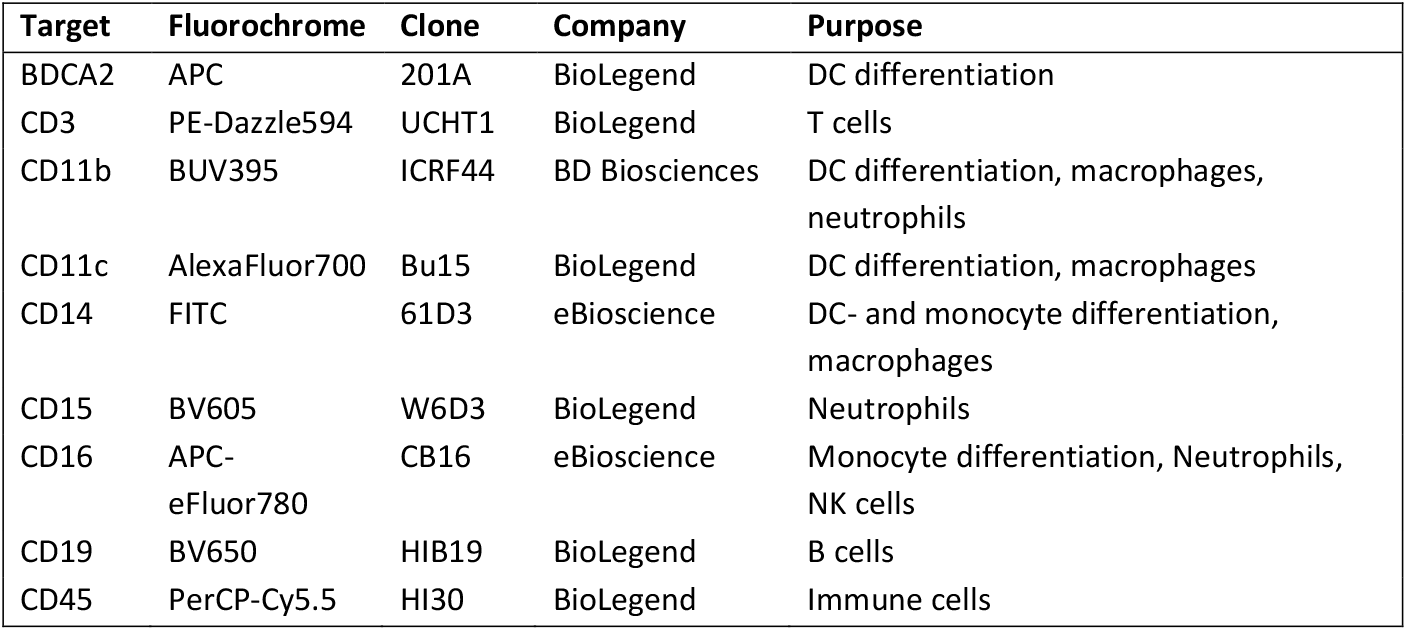
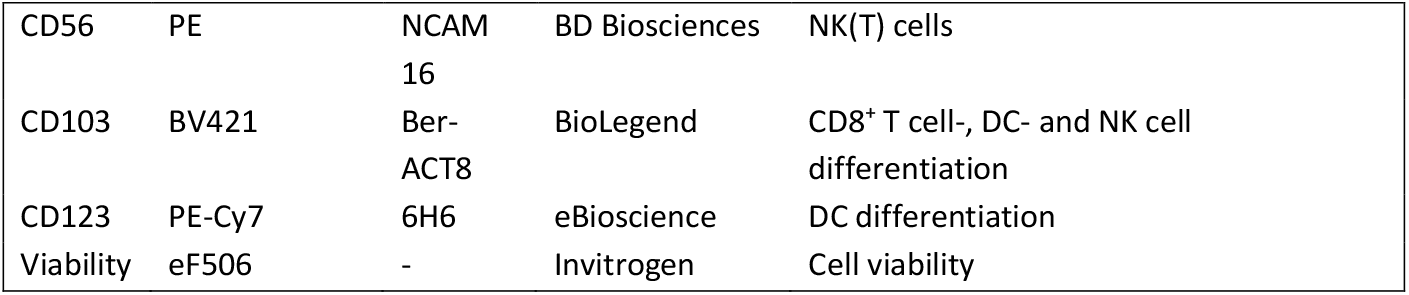
Flow cytometry panel for PBMC and placenta: Myeloid cells. CD = cluster of differentiation, DC = dendritic cell, NK cell = natural killer cell, BDCA2 = blood dendritic cell antigen 2. Viability is measured using fixable viability dye.

**Table 2.** Flow cytometry panel for PBMC and placenta: B cells. CD = cluster of differentiation, IgD = immunoglobulin D, IgM = immunoglobulin M. Viability is measured using fixable viability dye.

| Target | Fluorochrome | Clone | Company | Purpose |
| --- | --- | --- | --- | --- |
| CD19 | BV650 | HIB19 | BioLegend | B cells |
| CD21 | PE-Cy7 | B-ly4 | BD Biosciences | B cell differentiation |
| CD24 | BV421 | ML5 | BioLegend | B cell differentiation |
| CD27 | APC-Fire750 | O323 | BioLegend | B- and CD8<sup>+</sup> T cell differentiation |
| CD38 | PE | HIT2 | BioLegend | B cell differentiation |
| CD45 | PerCP-Cy5.5 | HI30 | BioLegend | Immune cells |
| CD138 | APC | MI15 | BioLegend | B cell differentiation |
| IgD | BV785 | IA6-2 | BioLegend | B cell differentiation |
| IgM | FITC | G20-127 | BD Biosciences | B cell differentiation |
| Viability | eF506 | - | Invitrogen | Cell viability |

**Table 3.** Flow cytometry panel for PBMC and placenta: T cells. CD = cluster of differentiation, TCRγδ = T cell receptor γδ. Viability is measured using fixable viability dye.

| Target | Fluorochrome | Clone | Company | Purpose |
| --- | --- | --- | --- | --- |
| CCR7 | PE-Cy7 | 3D12 | BD Biosciences | CD4<sup>+</sup>-, and CD8<sup>+</sup> T cell differentiation |
| CD3 | APC-R700 | UCHT1 | BD Biosciences | T cells |
| CD4 | BUV563 | SK3 | BD Biosciences | CD4<sup>+</sup> T cells |
| CD8 | BV785 | RPA-T8 | BioLegend | CD8<sup>+</sup> T cells |
| CD25 | APC | M-A251 | BioLegend | CD4<sup>+</sup> T cell differentiation |
| CD45 | PerCP-Cy5.5 | HI30 | BioLegend | Immune cells |
| CD45RA | eFluor450 | HI100 | eBioscience | $\gamma\delta$- CD4<sup>+</sup>-, CD8<sup>+</sup> T cell differentiation |
| CD45RO | PE | UCHL1 | BioLegend | $\gamma\delta$- CD4<sup>+</sup>-, CD8<sup>+</sup> T cell differentiation |
| CD56 | BV605 | HCD56 | BioLegend | $\gamma\delta$ differentiation, NK(T) cells |
| CD127 | APC-eFluor780 | eBioRDR5 | eBioscience | Non-NK ILC |
| TCR$\gamma\delta$ | FITC | B1 | BD Biosciences | $\gamma\delta$ T cells |
| Viability | eF506 | - | Invitrogen | Cell viability |

**Table 4.**
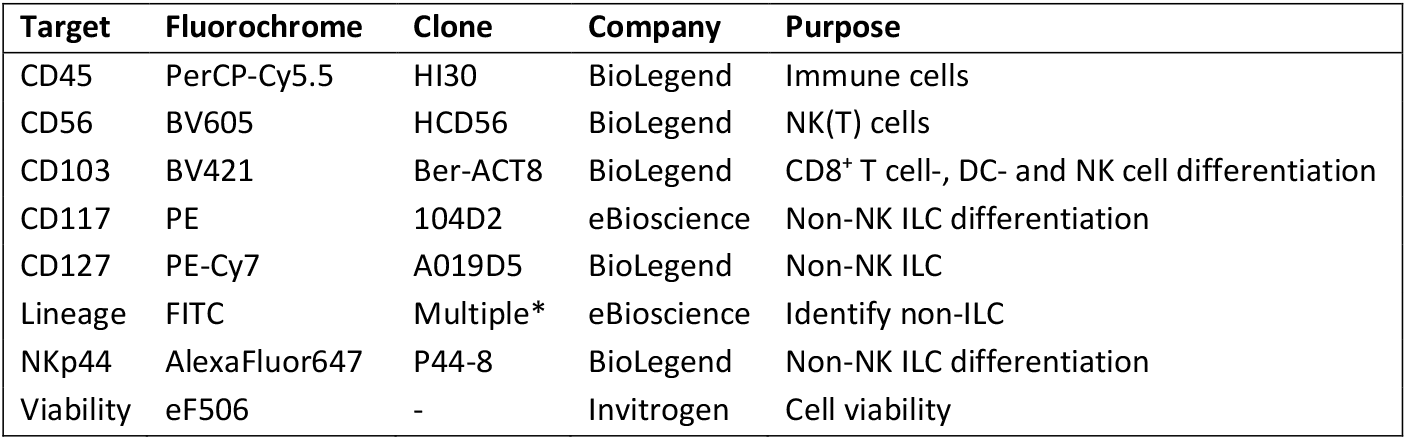
Flow cytometry panel for PBMC and placenta: innate lymphoid cells (ILC). CD = cluster of differentiation, *RPA-2.10, OKT3, 61D3, CB16, HIB19, TULY56, HIR2. Viability is measured using fixable viability dye.

**Table 5.** Flow cytometry panel for placenta: Non-immune cells. CD = cluster of differentiation, CK = Cytokeratin, HLA = human leukocyte antigen, * = intracellular staining. Viability is measured using fixable viability dye.

| Target | Fluorochrome | Clone | Company | Purpose |
| --- | --- | --- | --- | --- |
| CD31 | APC-eF780 | WM-59 | eBioscience | Endothelial cells |
| CD45 | PerCP-Cy5.5 | HI30 | BioLegend | Immune cells |
| CD235a | PE-Cy5 | HIR2 | BioLegend | Red blood cells |
| CK7* | AF647 | W16155A | BioLegend | Trophoblasts |
| HLA-DR | BV711 | L243 | BioLegend | Non-syncytiotrophoblasts |
| HLA-G | PE | MEM-G/9 | Exbio | Cytotrophoblasts, extravillous trophoblast, syncytiotrophoblasts |
| Viability | eF506 | - | Invitrogen | Cell viability |
| Vimentin* | AF488 | O91D3 | BioLegend | Decidual stromal cells, trophoblasts |

Whole blood is directly stained with the myeloid panel after which red blood cells are lysed using 1x RBC lysis buffer (BioLegend). PBMC and placenta cells are stained with the respective panels in PBA (PBS supplemented with 0.5% BSA, 0.01% NaN_3_), and then with fixable viability dye eF506 (Invitrogen, Carlsbad, CA, USA) in PBS. After staining, cells are fixed using Fix/Perm buffer (eBioscience, ThermoFisher Scientific). For the immune panels the cells are then measured at the LSR Fortessa (BD biosciences). For the non-immune panel, cells are stained with the intracellular antibodies in Perm buffer (eBioscience, ThermoFisher) and measured at the Sony SP6800. Data is analyzed using FlowJo v10.9.0.

### Statistical analysis

All statistical tests will be performed with a two-sided approach, using a 5% type I error threshold. Confidence intervals (CIs) will be set at 95%, unless otherwise indicated. Missing values will not be imputed. Collected data from patients lost to follow-up will be included in analysis where possible.

Variables will be reported as either means with standard error of the mean (SEM; normal distribution), medians with 95% CI (skewed distribution) or frequencies depending on the type and distribution of the data.

### Ethics and dissemination

This study is conducted according to the principles of the Declaration of Helsinki (version October 2013), and in accordance with the Medical Research Involving Human Subjects Act (WMO). This study was approved by the Medical Ethics Committee of Amsterdam UMC under registration number 2021.0774. Eligible women with SLE are given a verbal and written explanation of the study and the procedures involved, by their treating physician. Eligible healthy controls are asked by their treating physician whether they are interested in participating in the study, and subsequently are given verbal and written explanation of the study procedures by a member of the research team. Subjects who show interest in participation are contacted by a certified member of the research team. An independent physician is available to answer any question the subject may have. If the subject decides to participate written informed consent will be obtained before performance of any study-related activity.

Through identifying mechanisms underlying failing maternal-fetal tolerance, this study will identify potential therapeutic targets and biomarkers for development of pregnancy complications in women with SLE. In the long term, this may improve the development of preventive and therapeutic measures to reduce the incidence of pregnancy complications in SLE. Additionally, these findings may also be relevant to understand the pathogenesis of pregnancy complications in the general population.

The burden and risks of participation are negligible, considering the placenta is a waste product after child-birth and the blood collection will be performed simultaneously with routine clinical care as much as possible. There is no individual benefit from participation in this study.

Data will be disseminated through peer-reviewed publications in scientific journals, scientific presentations and through articles published in patient magazines and presentations for patient organizations.

Results from additional analyses performed on the stored biomaterials in a later stage, such as single-cell RNA sequencing, will be deposited in the appropriate data repositories (e.g. GEO Datasets).

## Data Availability

All data produced in the present study are available upon reasonable request to the authors

## AUTHOR CONTRIBUTIONS

WD, MB, LB and IB designed the study, with input on patient information and study setup from KC and DCRH. WD and JR wrote the manuscript. WD and APS designed the study protocols. SG and MG collect clinical data. All authors provided input on the manuscript and approved the final version.

## PATIENT AND PUBLIC INVOLVEMENT

Two patient research partners were involved in the design of the study, critically reviewed the patient information folders and contributed to this manuscript.

## ACKNOWLEDGEMENTS

We would like to thank the NVLE and Lupus Europe for their input and support for the FaMaLE study. In addition, we acknowledge the European Reference Network on Rare and Complex Connective Tissue Diseases (ERN ReCONNET) for declaring the Department of Rheumatology and Clinical Immunology of our centre as a member.

